# Multiscale connectomes reveal stage-dependent organization of cortical atrophy in Alzheimer’s disease

**DOI:** 10.64898/2026.04.16.26350695

**Authors:** Kexin Gao, Youjun Song, Jiahui Bao, Dezhong Yao, Bharat B. Biswal, Pan Wang

## Abstract

Cortical atrophy in Alzheimer’s disease (AD) follows a non-random spatial pattern, but whether different connectomes constrain atrophy across clinical stages and over time remains unclear. We investigated 726 participants in a discovery cohort and 563 in an independent validation cohort. The cohorts spanned the AD continuum, including the preclinical stage and individuals who converted during follow-up. We derived individual atrophy maps using normative modelling and assessed node-neighbor relationships across eight multiscale connectomes. Observed relationships were evaluated against three null models. Longitudinal analyses examined changes in node–neighbor relationships, while network diffusion models assessed how each connectome predicted follow-up atrophy. Cross-sectionally, no connectome exceeded all three null models in the preclinical groups. Structural connectivity, neurotransmitter receptor similarity, laminar similarity and sparse functional connectivity exceeded all three null models at later stages. Longitudinal increases in node–neighbor relationships were most widespread among mild cognitive impairment converters. Structural connectivity, neurotransmitter receptor similarity and gene co-expression generally provided better predictions of follow-up atrophy than other connectomes. Together, these findings show that structural connectivity and regional biological similarity increasingly shape cortical atrophy as AD progresses.

## 1. Introduction

Alzheimer’s disease (AD) is the most common cause of dementia and is defined biologically by the accumulation of amyloid β (Aβ) plaques and tau neurofibrillary tangles, followed by progressive neurodegeneration ^1,2^. Aβ deposition can begin more than a decade before symptom onset, yet clinical trajectories among Aβ-positive individuals vary considerably: some remain cognitively stable for many years, whereas others progress from normal cognition to mild cognitive impairment (MCI) and dementia. Gray matter atrophy measured with structural images is a proximal marker of neurodegeneration and tracks cognitive decline more closely than Aβ burden ^3^. Understanding the principles that govern where atrophy emerges and how it expands across the cortex is therefore central to explaining clinical heterogeneity and to predicting the disease course of individual patients.

Atrophy in AD is not randomly distributed. It follows a stereotyped spatial pattern that begins in the medial temporal lobe and extends to lateral temporal, posterior parietal and posterior cingulate cortex, regions that overlap closely with large scale intrinsic networks, particularly the default mode network ^4,5^. These observations gave rise to the network degeneration hypothesis, which proposes that pathology propagates along neural connections, with misfolded proteins spreading between connected neurons in a prion like manner ^6^. Consistent with this view, computational models in which pathology diffuses along the structural connectome reproduce the group-level pattern of atrophy in AD ^7,8^, and regions functionally connected to early affected epicenters are more likely to atrophy ^9^. Studies using tau positron emission tomography (PET) have further shown that tau accumulates preferentially in regions that are strongly connected to early affected regions ^10,11^. Network architecture also shape vulnerability through other routes, including the high metabolic demand of densely connected hubs ^12,13^ and shared gene expression and molecular composition among regions _14–16_.

Despite this progress, three questions remain open. First, previous studies have focused largely on individuals with MCI or dementia, leaving it unclear whether connectome architecture also constrains atrophy during the preclinical stage of AD. Second, most studies have examined a single type of connectivity, usually structural or functional connectivity, whereas the human cortex is organized by multiple, partly overlapping forms of interregional relationships spanning several spatial and biological scales, including gene co-expression, neurotransmitter receptor similarity, laminar similarity, metabolic connectivity and electrophysiological connectivity ^17^. Whether these connectomes differ in how strongly they constrain atrophy, and whether their relative contributions change along the AD continuum, is unknown. Third, most evidence comes from group-averaged data obtained at a single time point in individuals with dementia. It remains unclear whether network-constrained atrophy intensifies over time in individuals who progress clinically, and whether connectome-based models can predict future atrophy in individual patients ^18^.

Here, we investigated the relationships between cortical atrophy and multiple connectome architectures across Aβ-positive individuals at different clinical stages. Using normative modeling, we derived individual cortical atrophy maps relative to an Aβ-negative cognitively normal reference group. We then assessed whether regional atrophy was constrained by multiscale connectomes across structural, functional, molecular, metabolic and electrophysiological connectomes, evaluating these associations against spatial and connectome null models and a Euclidean-distance benchmark. Longitudinal analyses examined how these relationships changed within each clinical group. Finally, we used network diffusion models to identify candidate epicenters from baseline atrophy maps and compared how well different connectomes reproduced individual atrophy patterns at follow-up visits.

## 2. Results

We analyzed 726 participants in the Alzheimer’s Disease Neuroimaging Initiative (ADNI) as discovery cohort and 563 participants in Open Access Series of Imaging Studies (OASIS-3) as validation cohort. Details on subject inclusion are provided in the Methods (Supplementary Figure 1 and 2). Participants were classified into six groups according to amyloid-β (Aβ) status, baseline clinical diagnosis and subsequent diagnostic trajectory: Aβ-negative cognitively normal (Aβ− CN), Aβ-positive cognitively normal non-converters (Aβ+ CN-NC) and converters (Aβ+ CN-C), Aβ-positive MCI non-converters (Aβ+ MCI-NC) and converters (Aβ+ MCI-C), and Aβ-positive dementia (Aβ+ AD). Aβ− CN group were employed as healthy reference group and their demographic and clinical characteristics are provided in Supplementary Table 1.The mean diagnostic follow-up durations in the discovery cohort were 5.2 and 5.7 years for the Aβ+ CN-NC and Aβ+ CN-C groups, respectively, and 3.2 and 4.0 years for the Aβ+ MCI-NC and Aβ+ MCI-C groups, respectively. Details of diagnostic follow-up are provided in Supplementary Table 2.

The five Aβ+ groups in the discovery cohort did not differ significantly in age, sex, education, or handedness, with no significant differences observed (all *p* > 0.05; Table 1). The Clinical Dementia Rating (CDR) scores differed markedly across groups (*χ²* = 423.516, p < 0.001), reflecting increasing disease severity from Aβ+ CN to Aβ+ dementia. The Mini–Mental State Examination (MMSE) scores likewise declined across groups (*F* = 120.424, p < 0.001), with the lowest scores observed in the dementia Aβ+ group. The three Aβ+ groups in the validation cohort did not differ significantly in age, sex, education, and handedness, with no significant differences observed (all *p* > 0.05; Table 1).

**Table 1.** Demographic information.

|  |  | Discovery cohort: ADNI |  |  |  |  |  | Validation cohort: OASIS-3 |  |  |  |
|---|---|---|---|---|---|---|---|---|---|---|---|
|  |  | Aβ+<br>CN-NC<br>(n= 71) | Aβ+<br>CN-C<br>(n= 20) | Aβ+<br>MCI-NC<br>(n=76) | Aβ+<br>MCI-C<br>(n=48) | Aβ+<br>AD<br>(n=93) | Statistics | Aβ+-<br>CN-NC<br>(n=71) | Aβ+<br>CN-C<br>(n=18) | Aβ+<br>AD<br>(n=63) | Statistics |
| Age<br>(mean years ± SD) |  | 71.8±4.9 | 72.7±5.0 | 71.9±5.4 | 73.8±6.3 | 72.7±7.7 | F=0.986<br>P=0.416 | 70.6±4.5 | 70.6±5.5 | 71.9±5.3 | F=1.284<br>P=0.280 |
| Women<br>n(%) |  | 46(64.8%) | 11(55.0%) | 43(56.6%) | 22(45.8%) | 41(44.1%) | χ<sup>2</sup>= 8.338<br>P=0.080 | 49(69.0%) | 14(77.8%) | 39(61.9%) | χ<sup>2</sup>= 1.818<br>P=0.403 |
| Education years<br>(mean years ± SD) |  | 16.7±2.6 | 16.4±2.5 | 16.3±2.1 | 16.1±2.5 | 15.6±3.0 | F=2.006<br>P=0.094 | 16.0±2.4 | 15.8±2.4 | 15.6±2.2 | F=0.386<br>P=0.680 |
| Handedness,<br>n(%) | right | 66(93.0%) | 18(90.0%) | 66(86.8%) | 44(91.7%) | 87(93.5%) | χ<sup>2</sup>= 2.769<br>P= 0.597 | 62(87.3%) | 18(100%) | 55(87.3%) | χ<sup>2</sup>= 2.571<br>P= 0.276 |
|  | left | 5(7.0%) | 2(10.0%) | 10(13.2%) | 4(8.3%) | 6(6.5%) |  | 9(12.7%) | 0(0%) | 8(12.7%) |  |
| CDR<br>n(%) | 0 | 71(100%) | 20(100%) | 1(1.3%) | 0(0%) | 0(0%) | χ<sup>2</sup>= 423.516<br>P < 0.001 | 71(100%) | 17(94.4%) | 2(3.2%) | χ<sup>2</sup>= 140.132<br>P < 0.001 |
|  | 0.5 | 0(0%) | 0(0%) | 74(97.4%) | 48(100%) | 42(45.2%) |  | 0(0%) | 1(5.6%) | 45(71.4%) |  |
|  | 1 | 0(0%) | 0(0%) | 1(1.3%) | 0(0%) | 51(54.8%) |  | 0(0%) | 0(0%) | 16(25.4%) |  |
| MMSE scores<br>(mean ± SD) |  | 29.0±1.2 | 28.8±1.0 | 27.7±1.7 | 26.3±2.0 | 22.8±2.7 | F=120.424<br>P < 0.001 | 29.2±1.2 | 28.2±1.7 | 25.3±3.1 | F=50.194<br>P < 0.001 |

ADNI: Alzheimer’s Disease Neuroimaging Initiative; OASIS-3: Open Access Series of Imaging Studies; Aβ+ CN-NC: Aβ-positive cognitively normal non-converters; Aβ+ CN-C: Aβ-positive cognitively normal converters; Aβ+ MCI-NC: Aβ-positive mild cognitive impairment non-converters; Aβ+ MCI-C: Aβ-positive mild cognitive impairment converters; Aβ+ AD: Aβ-positive dementia; CDR: Clinical Dementia Rating; MMSE: Mini–Mental State Examination.

We first used normative modelling in Aβ− CN participants to derive individual maps of cortical atrophy from regional gray matter volume (GMV, Figure 1A). We then quantified node–neighbor relationships across eight connectomes, including structural connectivity (SC), gene co-expression (GC), neurotransmitter receptor similarity (RS), laminar similarity (LS), metabolic connectivity (MC), electrophysiological connectivity (EC), temporal similarity (TS) and functional connectivity (FC), and evaluated them against spatial and network null models and a Euclidean-distance benchmark. Longitudinal analyses tested whether these relationships changed over time (Figure 1B). Finally, we used network diffusion modelling to identify candidate epicenters from baseline atrophy maps and assessed how well simulations initiated from these regions reproduced individual follow-up atrophy patterns (Figure 1C).

**Figure 1.**
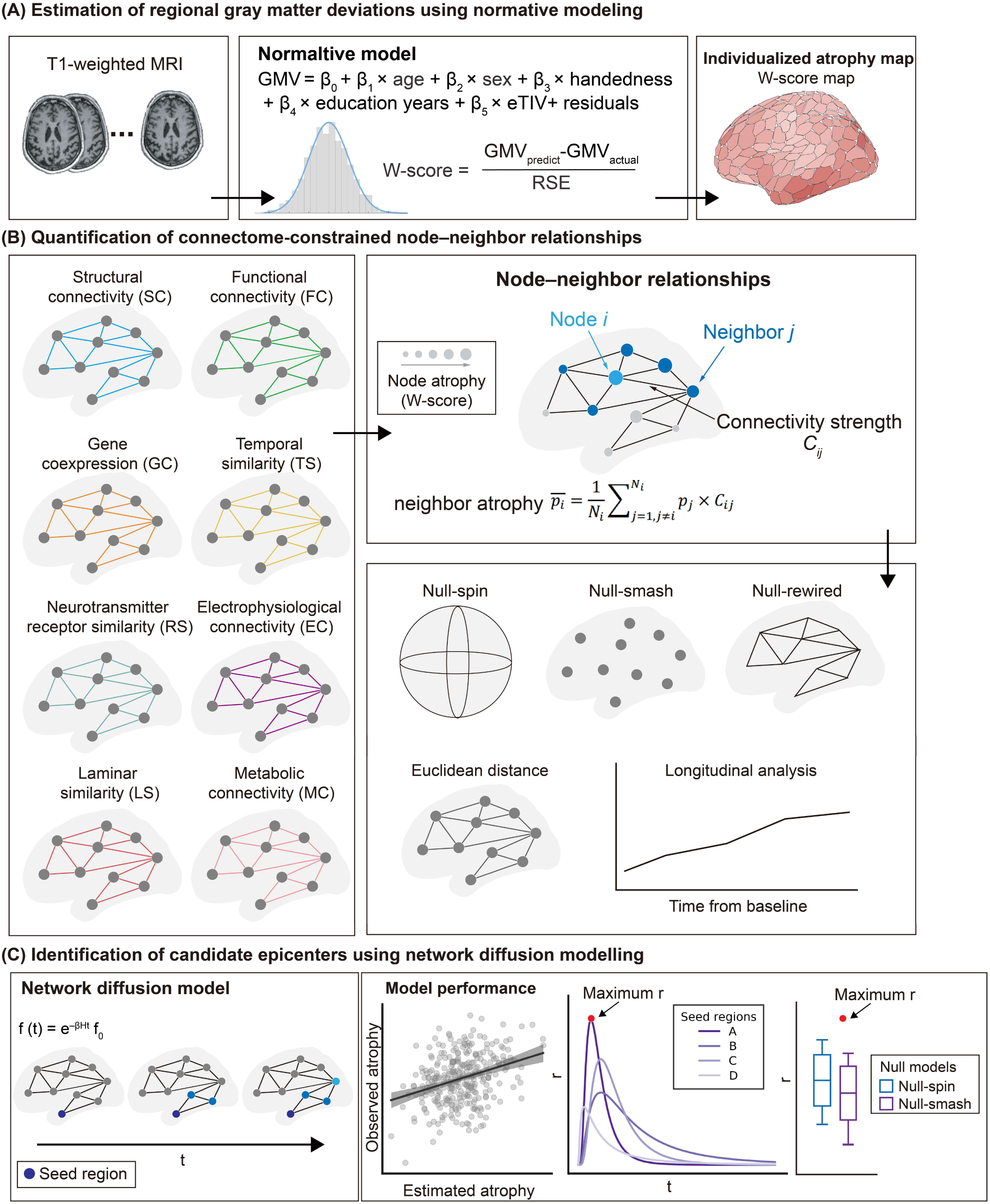
Overview of the study. (A) Normative modeling and individualized atrophy estimation. Regional gray matter volume (GMV) was modeled in Aβ− cognitively normal participants using a general linear model controlling for age, sex, handedness, education, and total intracranial volume (eTIV). Individualized W-scores were computed to quantify participant-specific deviations from normative expectations, generating maps of cortical atrophy. RSE: residual standard error. (B) Assessment of connectome-constrained atrophy. For each of the eight connectomes, we quantified the relationships between regional atrophy and the atrophy of connected neighbor regions. These node–neighbor relationships were then evaluated against two null models accounting for spatial autocorrelation (Null-spin and Null-smash), a rewired null model accounting for basic connectome topology (Null-rewire), and a benchmark based on Euclidean distance. Longitudinal changes in these relationships were also examined. (C) Identification of candidate epicenters and prediction of longitudinal atrophy progression. Each brain region was estimated in turn as a seed region, and a network diffusion model was used to estimate how atrophy might spread through the connectome over time. For each seed, model performance was quantified by the correlation (r) between estimated and observed regional atrophy across simulated time points. The highest correlation (maximum r) was retained for that seed, and its value was compared with distributions generated by spatially constrained null models (Null-spin and Null-smash). Regions with maximum r values exceeding a spatial autocorrelation null distributions were identified as candidate epicenters. The identified epicenters were then used to estimate longitudinal changes in regional atrophy.

### 2.1 Gray matter atrophy patterns and their association with cognition

Figure 2A shows group-average cortical atrophy maps, with more extensive and pronounced GMV loss across the Aβ+ CN, Aβ+ MCI and Aβ+ AD groups. Atrophy was relatively subtle in both Aβ+ CN groups and the Aβ+ MCI non-converter group, became more apparent in the Aβ+ MCI-C group, and was most widespread in the Aβ+ AD group. In the latter two groups, higher atrophy values were visible in lateral and medial temporal regions; the Aβ+ AD group also showed greater involvement of parietal and posterior medial regions. Across Aβ+ participants, greater global atrophy was associated with lower MMSE scores (r = −0.517, P < 0.001; Figure. 2B).

**Figure 2.**
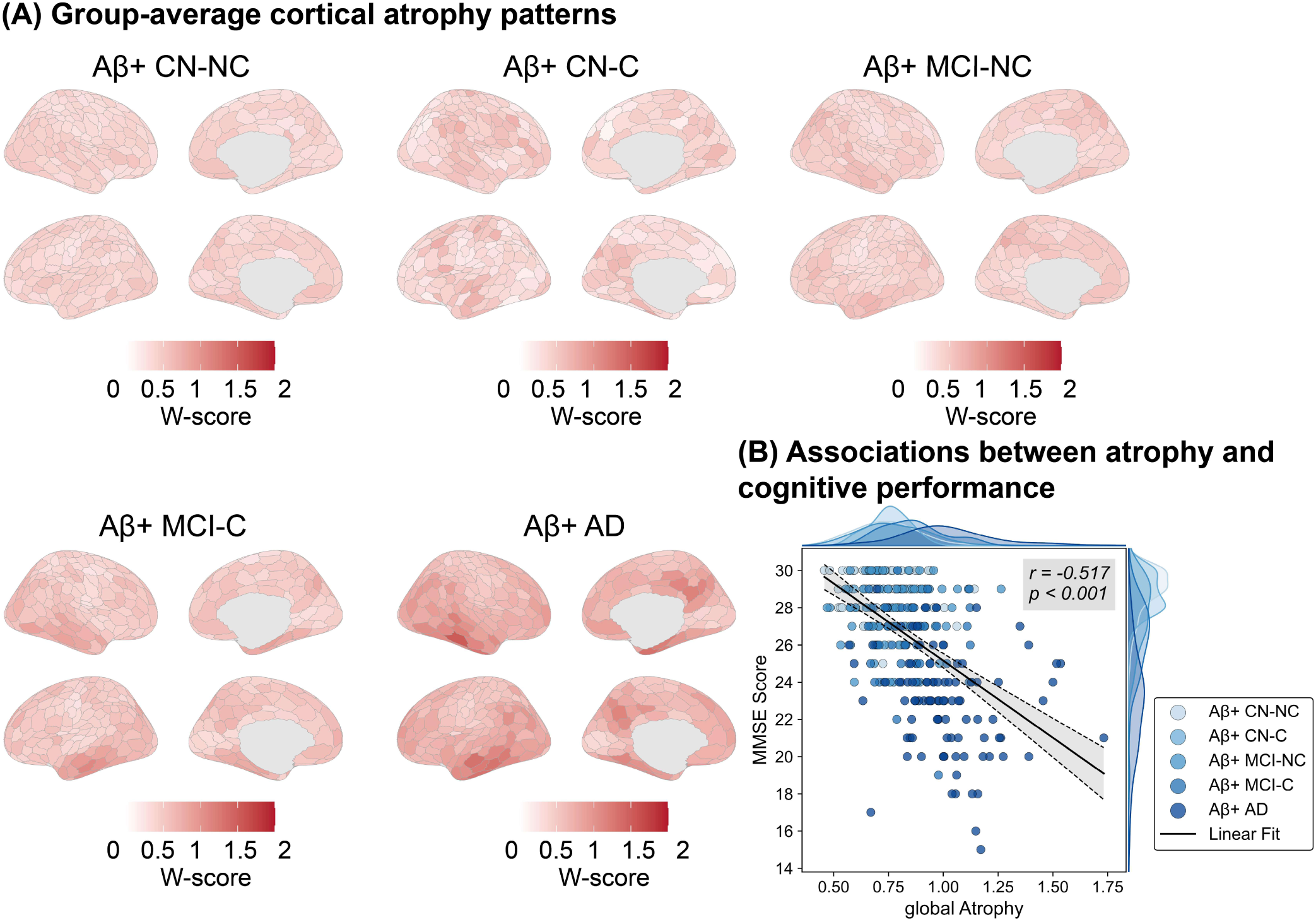
Group-average cortical atrophy patterns and their association with cognitive performance. (A) Cortical maps show mean regional atrophy W-scores in each group. (B) Association between global atrophy and Mini-Mental State Examination (MMSE) scores across all Aβ+ participants. Each point represents one participant, colored by group; marginal density curves show the distributions of global atrophy and MMSE scores.

### 2.2 Connectome-constrained atrophy across Aβ-positive groups

To assess whether brain connectome architecture constrains regional atrophy, we examined the relationship between atrophy in each region and the connectivity-weighted atrophy of its neighbors for each connectome, and further evaluated the observed group mean relationships against three null models (Figure 3). No connectome exceeded all three null models in either the Aβ+ CN-NC or Aβ+ CN-C group. The two measures bSC and wSC showed the strongest node–neighbor relationships across all five Aβ-positive groups, and both exceeded all three null models in the Aβ+ MCI-NC, Aβ+ MCI-C and Aβ+ AD groups. RS and LS exceeded all three null models only in the Aβ+ MCI-C and Aβ+ AD groups; GC, EC and TS did not do so in any group. In Aβ+ MCI-NC, FC20 and FC10 exceeded all three null models; this extended to FC30–FC5 in Aβ+ MCI-C and FC40–FC5 in Aβ+ AD.

**Figure 3.**
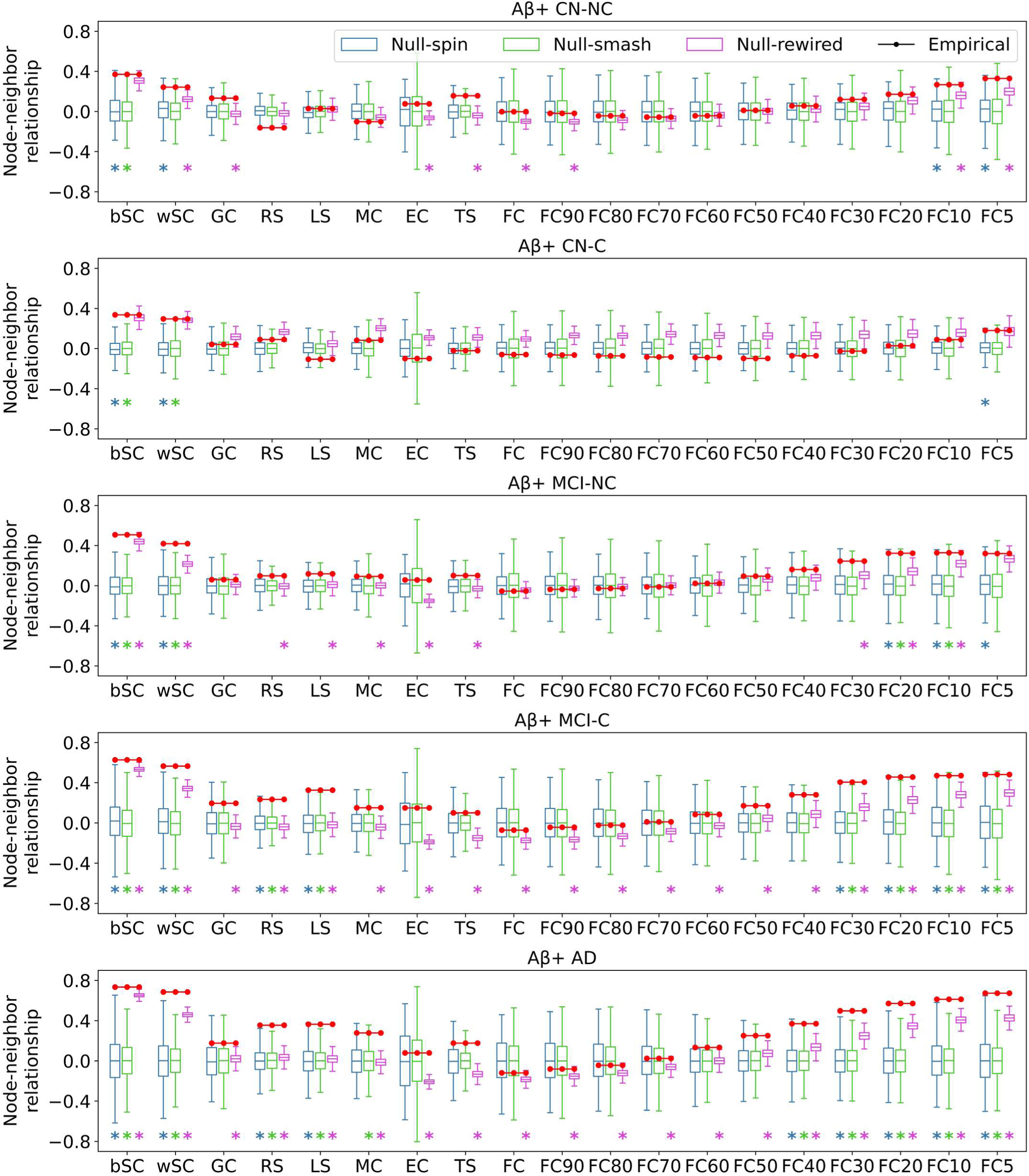
Node–neighbor relationships across amyloid-β-positive clinical groups and network null models. Red dots represent empirical node–neighbor relationships for each group. Colored asterisks indicate significant differences between the empirical relationship and the corresponding null distribution after FDR correction (*p*_FDR <_ 0.05). Aβ+ CN-NC: Aβ-positive cognitively normal non-converters; Aβ+ CN-C: Aβ-positive cognitively normal converters; Aβ+ MCI-NC: Aβ-positive mild cognitive impairment non-converters; Aβ+ MCI-C: Aβ-positive mild cognitive impairment converters; Aβ+ AD: Aβ-positive dementia; bSC: binary structural connectivity; wSC: weighted structural connectivity; GC: gene co-expression; RS: neurotransmitter receptor similarity; LS: laminar similarity; MC: metabolic connectivity; EC: electrophysiological connectivity; TS: temporal similarity; FC: functional connectivity. FC90–FC5: retaining the strongest 90%–5% of FC connections.

### 2.3 Connectome-based node–neighbor relationships exceed those explained by spatial proximity

Since neighboring regions are often both strongly connected and similarly affected, we next asked whether these relationships exceeded those expected from spatial proximity alone, by comparing individual node–neighbor relationships for each connectome with those obtained from the Euclidean distance network (Figure 4A). Across all five Aβ-positive groups, bSC and wSC exhibited significantly higher node–neighbor relationships than those in the Euclidean distance network. Among functional networks, FC5 showed significantly higher relationships in all groups, extending to FC10 and FC20 in the Aβ+ CN-NC, Aβ+ MCI-NC and Aβ+ MCI-C groups, and to FC30 and FC40 in the Aβ+ AD group. RS and MC also exhibited significantly higher relationships in all groups except Aβ+ CN-C. By contrast, GC, LS, EC, TS, and the denser functional networks (FC through FC50) did not exceed the Euclidean distance network in any group.

**Figure 4.**
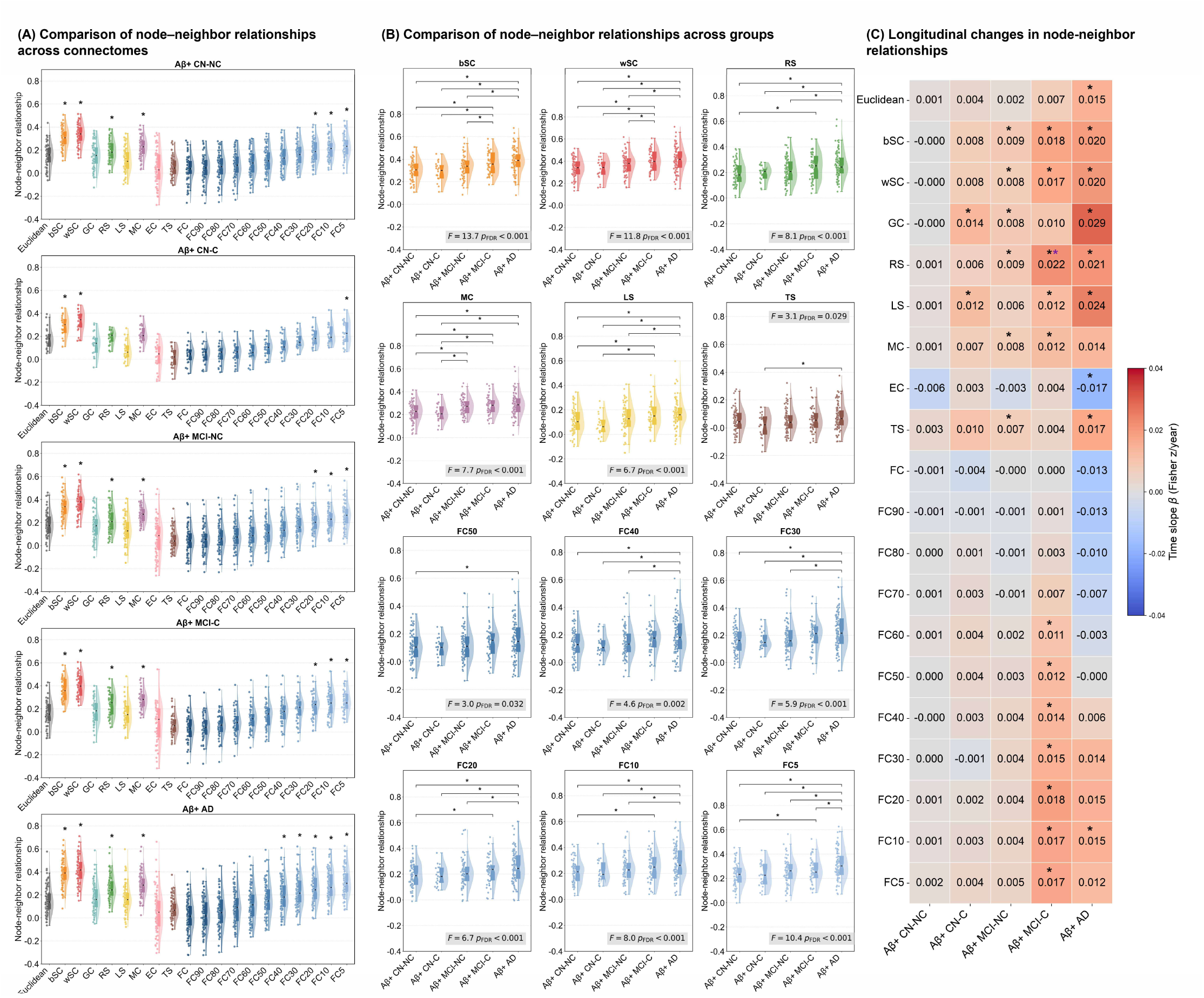
Connectome-specific node–neighbor relationships and their longitudinal changes across Aβ-positive groups. (A) Distributions of node–neighbor relationships across connectomes and the Euclidean-distance benchmark in each Aβ-positive group. Points represent individual participants; violin plots show the corresponding distributions. Asterisks indicate connectomes for which the node–neighbor relationship was significantly higher than that of the Euclidean-distance benchmark (*P*_FDR_ < 0.05). (B) Comparisons of node–neighbor relationships across groups. Statistical results from analysis of covariance (ANCOVA) are displayed within each panel. (C) Estimated annual change in Fisher *z*-transformed node–neighbor relationships for each connectome and group. Cell values give the time slope (β, Fisher *z* per year). Black asterisks indicate significant time effect, and purple asterisks indicate time slopes significantly higher than those for Euclidean proximity (*P_FDR_* < 0.05). Aβ+ CN-NC: Aβ-positive cognitively normal non-converters; Aβ+ CN-C: Aβ-positive cognitively normal converters; Aβ+ MCI-NC: Aβ-positive mild cognitive impairment non-converters; Aβ+ MCI-C: Aβ-positive mild cognitive impairment converters; Aβ+ AD: Aβ-positive dementia; bSC: binary structural connectivity; wSC: weighted structural connectivity; GC: gene co-expression; RS: neurotransmitter receptor similarity; LS: laminar similarity; MC: metabolic connectivity; EC: electrophysiological connectivity; TS: temporal similarity; FC: functional connectivity. FC90–FC5: retaining the strongest 90%–5% of FC connections.

Node–neighbor relationships differed across the five Aβ-positive groups for bSC, wSC, RS, MC, LS, TS and the FC50-FC5 connectomes shown in Figure 4B (*P*_FDR_ < 0.05). For most of these connectomes, relationships were generally higher in the Aβ+ MCI-C and Aβ+ AD groups than that in the Aβ+ CN groups, although the significant pairwise differences varied by connectome. The largest group effects were observed for bSC (*F* = 13.7) and wSC (*F* = 11.8; both *P*_FDR <_ 0.001). Node–neighbor relationships for EC, GC, FC, and FC90-FC60, as well as the Euclidean-distance benchmark, did not differ significantly across the five Aβ-positive groups (all *P*_FDR_ > 0.05; Supplementary Figure 3).

The estimated time effect varied by connectome and clinical group (Figure 4C). Longitudinal analyses included 71 participants with 202 scans in the Aβ+ CN-NC group, 20 with 82 scans in the Aβ+ CN-C group, 76 with 254 scans in the Aβ+ MCI-NC group, 48 with 178 scans in the Aβ+ MCI-C group, and 93 with 211 scans in the Aβ+ AD group. No connectome showed a significant time effect in the Aβ+ CN-NC group. In the Aβ+ CN-C group, positive time effects were observed for GC and LS. In the Aβ+ MCI-NC group, bSC, wSC, GC, RS, MC and TS showed positive time effects. The Aβ+ MCI-C group showed the broadest pattern of increases, including bSC, wSC, RS, LS, MC and FC60–FC5. Among these, only the RS time slope was significantly greater than the slope for Euclidean proximity (*P*_FDR <_ 0.05). In the Aβ+ AD group, positive time effects were observed for bSC, wSC, GC, RS, LS, TS and FC10, whereas EC showed a negative time effect. Overall, significant longitudinal increases in node–neighbor relationships were most widespread in the Aβ+ MCI-C group.

### 2.4 Spatial distribution of candidate epicenters identified from baseline data

Using baseline atrophy maps from all participants, the spatial distribution of candidate epicenters differed across connectomes (Figure 5). Epicenters identified using bSC, wSC, RS and LS were more concentrated in lateral temporal regions. MC and EC identified relatively few epicenters. Across FC connectomes, epicenters were concentrated in temporal and posterior cortical regions, with these patterns becoming more prominent as weaker connections were removed. The number of candidate epicenters per participant across different connectomes was shown in the Supplementary Figure 4.

**Figure 5.**
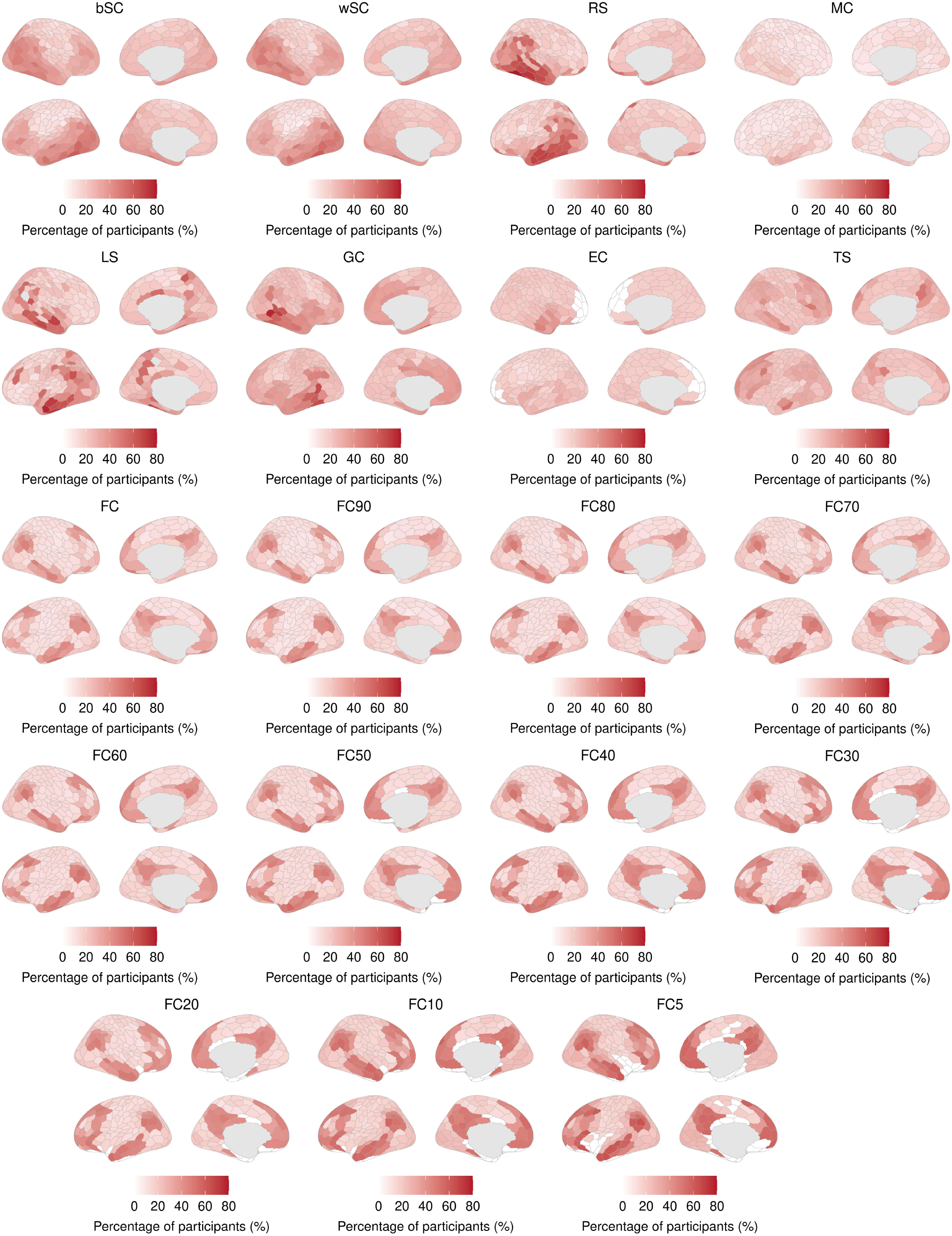
Spatial distribution of candidate epicenters identified from baseline data. Candidate epicenters were identified for each participant using a network diffusion model (NDM) applied separately to each connectome. Cortical maps show the percentage of participants for whom each region was identified as a candidate epicenter. bSC: binary structural connectivity; wSC: weighted structural connectivity; GC: gene co-expression; RS: neurotransmitter receptor similarity; LS: laminar similarity; MC: metabolic connectivity; EC: electrophysiological connectivity; TS: temporal similarity; FC: functional connectivity. FC90–FC5: retaining the strongest 90%–5% of FC connections.

### 2.5 Longitudinal prediction of cortical atrophy across connectomes

The NDM prediction performance differed across connectomes and follow-up intervals (Figure 6). At 6 months, wSC, RS and GC outperformed selected other connectomes, whereas significant advantages for bSC were less widespread. At 12 months, wSC, bSC, RS and GC showed more consistent advantages over several other connectomes; wSC also slightly outperformed bSC. Among FC variants, FC10 and FC5 outperformed several denser networks. At 24 months, RS and GC showed the clearest advantages, outperforming many FC variants without differing significantly from wSC or bSC. At 36 months, RS and GC remained numerically among the stronger connectomes, although fewer pairwise differences were significant and several comparisons could not be evaluated. At this interval, FC5 outperformed several denser FC variants. Overall, SC, RS and GC connectomes generally provided stronger fits to follow-up atrophy patterns, while FC10 and FC5 outperformed several denser FC variants. Repeating the comparisons using a fixed number of top-ranked candidate epicenters per connectome yielded a similar overall pattern: SC, RS and GC generally provided stronger fits to follow-up atrophy maps, and sparse FC variants outperformed several denser variants (Supplementary Figure 5).

**Figure 6.**
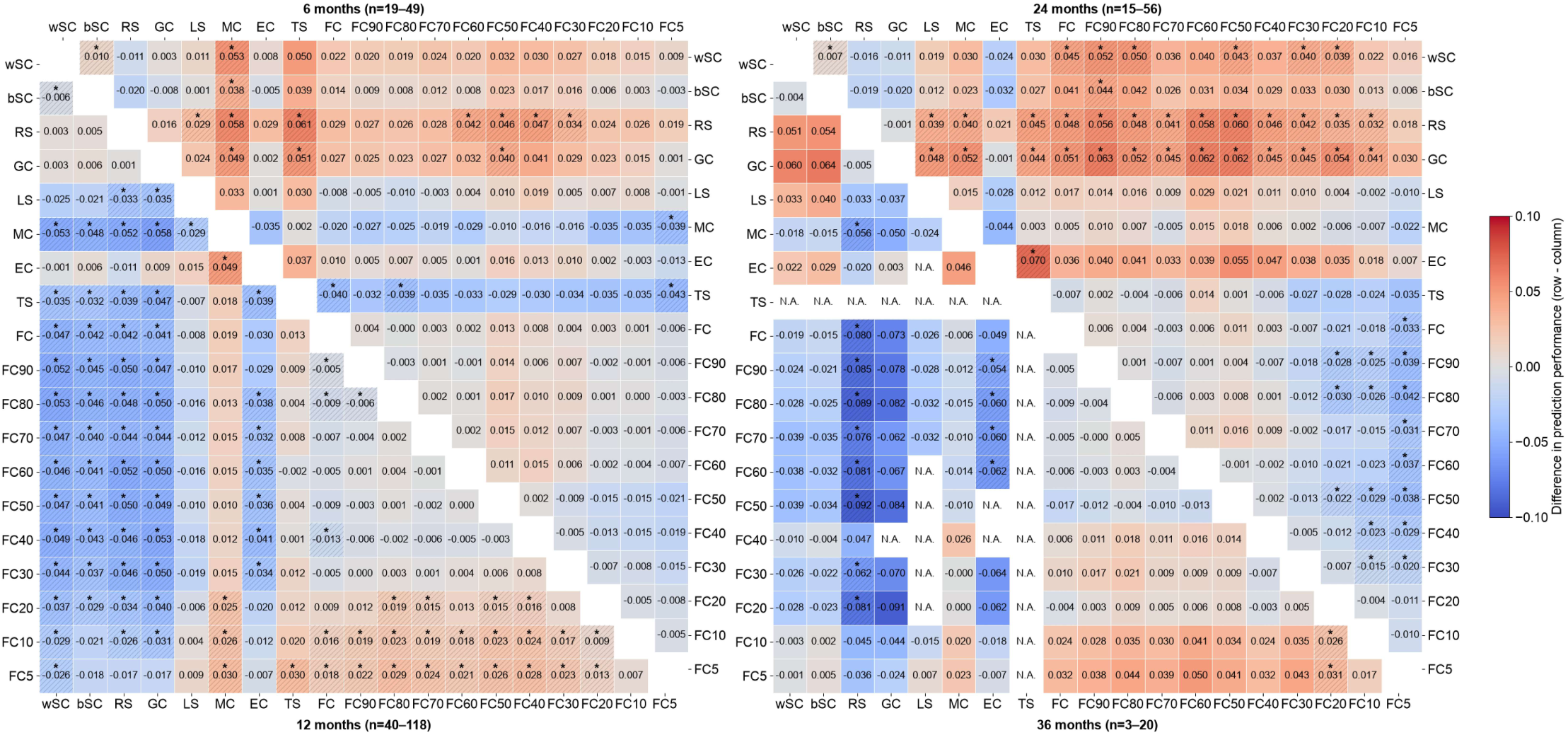
Pairwise comparisons of network diffusion model (NDM) prediction performance at 6, 12, 24 and 36 months. Cell values represent the mean difference in NDM prediction performance between the connectome indicated by the row and column labels. Asterisks and diagonal hatching mark significant differences (*P*_FDR <_ 0.05). The *n* range shown for each follow-up interval indicates the minimum and maximum numbers of participants with prediction estimates for both connectomes in a pairwise comparison. Each participant contributed at most one visit within the ±2-month window around the target month; comparisons with fewer than 10 paired participants are shown as N.A. bSC: binary structural connectivity; wSC: weighted structural connectivity; GC: gene co-expression; RS: neurotransmitter receptor similarity; LS: laminar similarity; MC: metabolic connectivity; EC: electrophysiological connectivity; TS: temporal similarity; FC: functional connectivity. FC90–FC5: retaining the strongest 90%–5% of FC connections.

### 2.6 External validation of connectome-constrained cortical atrophy

The independent OASIS-3 validation cohort reproduced the pattern of connectome-constrained atrophy observed in the discovery cohort (Figure 7). Consistent with the discovery findings, no connectome exceeded all three null models in either Aβ+ CN group. In Aβ+ AD, wSC, RS, LS and FC30–FC5 exceeded all three null models, whereas bSC exceeded the two spatial null models but not the rewired null model (Figure 7A).

**Figure 7.**
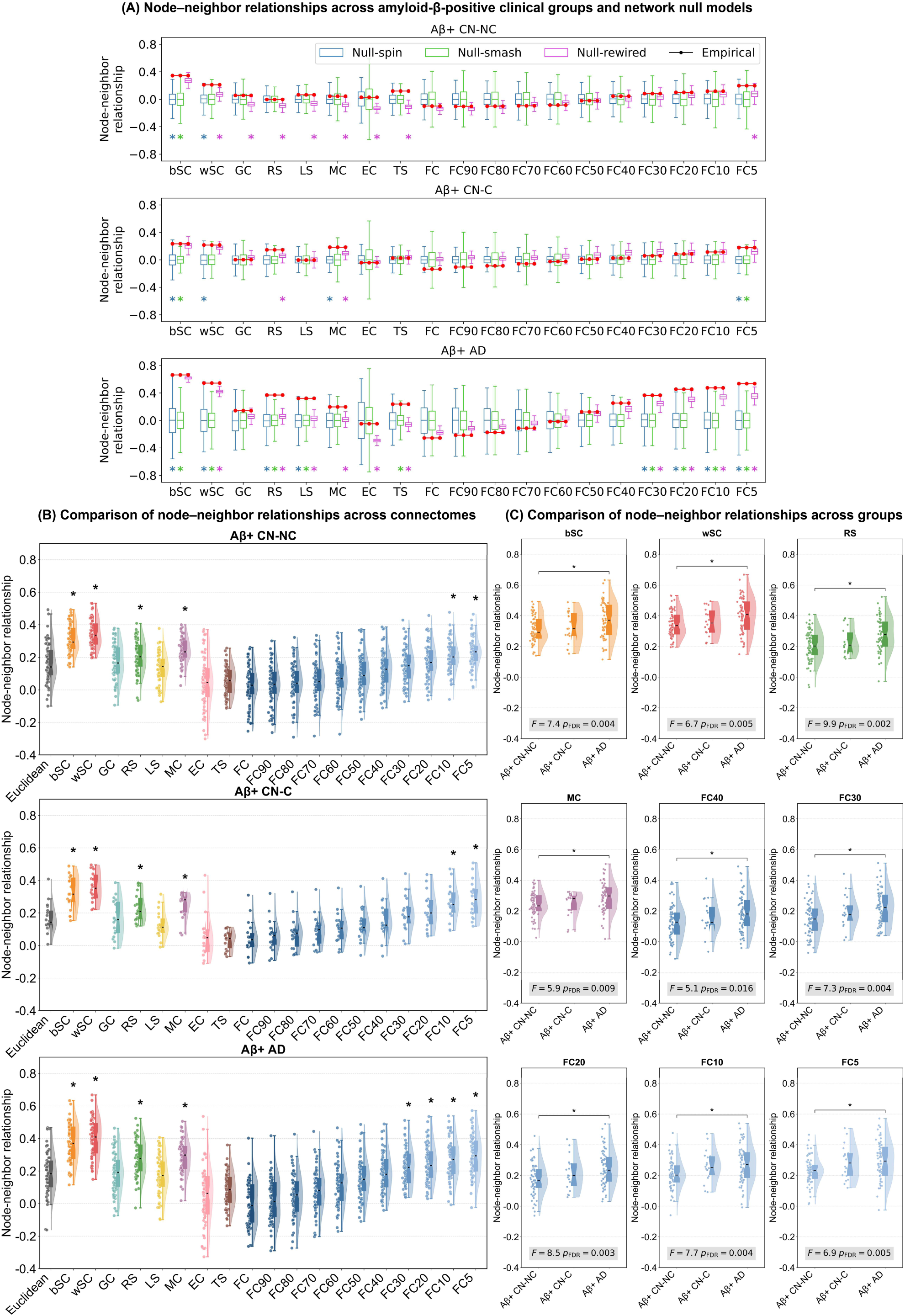
External validation of connectome-constrained cortical atrophy. (A) Node–neighbor relationships across amyloid-β-positive clinical groups and network null models. Red dots represent empirical node–neighbor relationships for each group. Colored asterisks indicate significant differences between the empirical relationship and the corresponding null distribution after FDR correction (*p*_FDR_ < 0.05). (B) Distributions of node–neighbor relationships across connectomes and the Euclidean-distance benchmark in each Aβ-positive group. Points represent individual participants; violin plots show the corresponding distributions. Asterisks indicate connectomes for which the node–neighbor relationship was significantly higher than that of the Euclidean-distance benchmark (*P*_FDR_ < 0.05). (C) Comparisons of node–neighbor relationships across groups. Statistical results from analysis of covariance (ANCOVA) are displayed within each panel. Aβ+ CN-NC: Aβ-positive cognitively normal non-converters; Aβ+ CN-C: Aβ-positive cognitively normal converters; Aβ+ AD: Aβ-positive dementia; bSC: binary structural connectivity; wSC: weighted structural connectivity; GC: gene co-expression; RS: neurotransmitter receptor similarity; LS: laminar similarity; MC: metabolic connectivity; EC: electrophysiological connectivity; TS: temporal similarity; FC: functional connectivity. FC90–FC5: retaining the strongest 90%–5% of FC connections.

At the individual level, bSC, wSC, RS, MC, FC10 and FC5 showed significantly higher node–neighbor relationships than the Euclidean-distance benchmark in all three validation groups; FC30 and FC20 additionally exceeded this benchmark in Aβ+ AD (Figure 7B). These findings broadly agreed with those in the discovery cohort, although the advantages of RS and MC were also significant in Aβ+ CN-C in the validation cohort. Between-group differences were significant for bSC, wSC, RS, MC and FC40–FC5 (all P < 0.05), with post hoc comparisons showing higher relationships in Aβ+ AD than in Aβ+ CN-NC (Figure 7C). Together, these results support the reproducibility of stronger connectome-constrained cortical atrophy in Aβ+ AD than in cognitively normal individuals.

## 3. Discussion

In this study, we compared how cortical atrophy relates to multiple connectomes across the Aβ-positive clinical continuum. First, neither the Aβ+ CN-NC nor the Aβ+ CN-C showed significant node–neighbor relationships, suggesting that atrophy patterns in the early stages of AD were not constrained by brain connectome architecture. Second, as AD progressed to later clinical stages, cortical atrophy became increasingly constrained by bSC, wSC, RS, LS, MC and sparse FC, with these constraints strengthening over time. Finally, wSC, bSC, RS, GC and sparse FC generally provided better predictions of individual follow-up atrophy patterns than other connectomes. Together, these findings support a model in which early degeneration shows limited dependence on connectome architecture, whereas later-stage degeneration becomes increasingly organized by specific biological connectome constraints.

### Preclinical atrophy: network vulnerability before widespread cortical degeneration

In the Aβ+ CN groups, cortical atrophy was subtle and no connectome showed significant node–neighbor relationships that exceeded all three null models. This apparent dissociation may reflect a temporal gap between the emergence of connectome-organized pathology and the development of measurable cortical volume loss. Aβ accumulation has been observed early within the default mode network alongside changes in brain connectivity ^19^. Connectome-based modeling of tau-PET data has further shown that tau follows neuronal communication pathways even in cognitively unimpaired individuals, with this pattern amplified in the presence of Aβ ^11^. These observations suggest that network-organized pathology may precede a robust network-constrained pattern of cortical atrophy, consistent with our finding that, no connectome exceeded all three null models in either Aβ+ CN group. The longitudinal distinction between the two CN groups further supports this interpretation: no connectome showed a significant time effect in Aβ+ CN-NC, whereas GC and LS increased in Aβ+ CN-C. Thus, the transition towards clinical impairment may involve an early shift from anatomically patterned vulnerability to a more detectable relationship between atrophy and the brain connectome architecture.

### Structural routes, regional similarity and the functional backbone of atrophy

SC and regional similarity appear to capture complementary components of cortical degeneration. bSC and wSC showed the strongest cross-sectional node–neighbor relationships, consistent with the view that anatomical connections provide routes along which disease-related effects become coordinated ^7,8,20^. Yet SC-defined candidate epicenters were broadly distributed, whereas RS and several other similarity-based connectomes produced more spatially concentrated patterns. Moreover, RS and GC, alongside SC, provided comparatively strong predictions of follow-up atrophy maps. These results support a model in which SC specifies the routes available for disease-related change, while regional molecular similarity helps determine where that change is most strongly expressed ^21^. This interpretation is consistent with evidence that selective vulnerability depends on the molecular properties of cortical regions ^22^ and that gene-expression gradients can modify the spatial pattern of tau pathology ^23^. The RS finding is particularly relevant because neurotransmitter systems influence regional excitability and synaptic function; the cholinergic system, for example, is affected in AD and has widespread cortical projections ^24^. However, RS combines multiple receptor and transporter systems, so the present results identify a neurochemical axis of vulnerability rather than a specific transmitter mechanism.

FC showed a different dependence on network density. Stronger, sparser FC variants were more consistently related to atrophy than dense FC matrices, particularly in the MCI and AD groups. Structural pathways contribute to the organization of functional connectivity ^25^, providing one reason why strong FC edges may retain disease-relevant information while many weak correlations dilute it. The increasing range of FC densities that exceeded all three null models—from FC20 and FC10 in MCI-NC to FC30–FC5 in MCI-C and FC40–FC5 in AD—also resonates with models of cascading network failure, in which disruption of initially vulnerable systems is followed by broader network involvement ^26^. Our results extend that account by showing that the relationship between FC and cortical atrophy depends on both clinical stage and the strength of the functional connections retained.

### Connectome constraint strengthens with clinical progression and informs longitudinal atrophy patterns

The longitudinal analysis provides a view of connectome organization that cannot be inferred from cross-sectional group differences alone. Positive time effects were most widespread in Aβ+ MCI-C, involving SC, RS, LS, MC and sparse FC, while additional effects were present in MCI-NC and AD. Of particular interest, the increase in RS-based node–neighbor relationships in MCI-C exceeded that expected from Euclidean proximity. This suggests that, during conversion from MCI to dementia, the developing atrophy pattern becomes increasingly aligned with cortical neurochemical organization. Such a pattern is compatible with evidence that the distribution of tau pathology depends on both network connections and regional molecular properties ^23^. It also helps explain why a purely anatomical account may be insufficient ^18^: connected regions provide potential routes of involvement, but their biological characteristics may influence the degree to which degeneration develops along those routes.

The NDM results reinforce this multilevel account. wSC, bSC, RS and GC generally provided stronger fits to individual follow-up atrophy maps than many other connectomes, although their relative performance varied by interval. Because follow-up atrophy maps are strongly correlated with baseline maps, part of this performance reflects persistence of the baseline atrophy pattern rather than prediction of newly emerging change. The contribution of RS and GC is notable because neither represents a direct physical pathway. Their performance suggests that models of disease progression gain information not only from which regions are connected, but also from which regions share properties that make them vulnerable ^22,23^. Cortical geometry is also an important determinant of large-scale brain organization ^27,28^; the interpretation of a connectome-based model therefore depends on whether it improves upon spatial benchmarks. Taken together, the cross-sectional, longitudinal and NDM findings support a progression from relatively limited connectome-specific cortical atrophy in the preclinical stage towards degeneration organized jointly by anatomical connections and regional biological similarity in AD. These findings motivate models that combine connectivity, individual epicenters and regional vulnerability when forecasting disease evolution ^29^.

### Limitations

Several limitations should be considered. First, the connectomes were derived from healthy young adults and postmortem or atlas data and were applied as group templates. This allowed many modalities to be compared within a common parcellation, but it does not capture age related or disease related changes in connectivity, nor individual variation; patient specific connectomes may improve both the sensitivity of the node–neighbor analysis and the accuracy of diffusion models. Second, the analyses were restricted to the cortex because the connectomes were defined on the Schaefer parcellation, so the hippocampus and amygdala, which are among the earliest and most severely affected structures in AD, were not modeled. Third, some samples were small, particularly the Aβ+ CN-C group (n = 20) and the 36 month follow up (n = 6-22), which limits statistical power and may partly explain the weaker findings in converting CN Aβ+ individuals. Fourth, conversion status depends on the length of follow up, and some participants classified as non-converter may convert later, which would attenuate differences between non-converter and converter groups. Fifth, we did not model tau pathology, which is more closely related to neurodegeneration than Aβ; future work should test whether network constrained atrophy is mediated by tau spreading along the same connectomes. Sixth, we analyzed only regional volume loss relative to the normative model, and diffusion models used a fixed diffusion constant and seeds selected at baseline; alternative parameter choices may change absolute model performance.

## Conclusions

Our findings reveal a stage-dependent organization of cortical atrophy across the Aβ-positive AD continuum. In cognitively normal individuals, atrophy shows limited connectome-specific organization; after the onset of cognitive impairment, its spatial pattern becomes strongly constrained by structural connectivity and increasingly aligned with neurotransmitter receptor, laminar and sparse functional connectomes. These relationships strengthen most broadly during progression from MCI to dementia. Structural connectivity provides a scaffold for the distribution of atrophy, while shared molecular properties help explain which regions become most affected. The ability of structural, neurotransmitter receptor and gene co-expression connectomes to predict individual follow-up atrophy patterns further shows that no single connectome captures the full course of degeneration. Together, these results establish a multilevel framework for understanding how cortical atrophy becomes organized as AD progresses and identify the transition from MCI to dementia as a critical period of connectome-related change.

## 4. Methods

### 4.1 Participants

#### 4.1.1 Discovery cohort

A total of 3,254 participants were obtained from ADNI database (adni.loni.usc.edu). 2,528 participants were excluded for the reasons shown in Supplementary Figure 1, resulting in 726 participants ultimately being retained for this study. The ADNI study was approved by the institutional review boards of all participating sites. Written informed consent was obtained from all participants or their authorized representatives. The study was performed in accordance with the ethical standards as laid down in the 1964 Declaration of Helsinki and its later amendments or comparable ethical standards. Inclusion criteria required the availability of amyloid-PET, T1-weighted magnetic resonance imaging (MRI), and demographic data. All baseline imaging modalities were obtained within a 1-year window. Aβ status was determined based on established tracer-specific standardized uptake value ratios (SUVR) cut-points: ¹⁸F-florbetapir (AV45) > 1.11, ¹⁸F-florbetaben > 1.08, and ¹⁸F-flutemetamol > 1.14 ^30–32^. Participants were classified into six groups according to Aβ status, baseline clinical diagnosis and subsequent diagnostic trajectory: Aβ− CN, Aβ+ CN-NC, Aβ+ CN-C, Aβ+ MCI-NC, Aβ+ MCI-C, and Aβ+ AD. Participants in the Aβ− CN and Aβ+ AD groups were required to retain their baseline diagnosis throughout diagnostic follow-up. Participants with Aβ+ CN or Aβ+ MCI at baseline were required to have at least 1 year of diagnostic follow-up. Among those with Aβ+ CN at baseline, converters were defined as participants who progressed to MCI or dementia during follow-up; among those with Aβ+ MCI at baseline, converters were defined as participants who progressed to dementia. Participants who did not meet these criteria were classified as non-converters.

#### 4.1.2 Validation cohort

External validation was performed in an independent cohort from OASIS-3 ^33^. From the OASIS-3 cohort, 1,335 participants were initially screened. After applying predefined inclusion and exclusion criteria, 563 participants were included (Supplementary Figure 2). Aβ status was determined from the mean SUVRs across the precuneus, prefrontal cortex, gyrus rectus and lateral temporal regions using the cerebellar cortex as the reference region. Aβ+ was defined by tracer-specific cut-offs: for ¹¹C-Pittsburgh compound B > 1.31 or FBP > 1.24 ^34^. Participants were assigned to the same six groups used in the discovery cohort. However, the Aβ+ MCI-NC and Aβ+ MCI-C groups were excluded from the external validation analyses because each contained fewer than five participants. All procedures were approved by the institutional review boards of participating sites, and written informed consent was obtained from all participants.

### 4.2 Neuroimaging acquisition and processing

Acquisition parameters for amyloid-PET scans followed the standardized ADNI protocols and T1-weighted MRI scans were acquired according to ADNI MRI scanner protocols (https://adni.loni.usc.edu/data-samples/adni-data/neuroimaging/mri/mri-scanner-protocols/). The T1-weighted anatomical MRI data were processed using the automated recon-all pipeline implemented in FreeSurfer (https://surfer.nmr.mgh.harvard.edu/fswiki/recon-all), which includes skull stripping, tissue segmentation, and cortical surface reconstruction. Regional gray matter volumes (GMV) were extracted using the Schaefer atlas with 400 cortical regions ^35^. To minimize site-related variability across scanners, ComBat harmonization ^36^ was applied to regional GMV measures with imaging site treated as a batch variable, while age, sex, education years, handedness and group were included as covariates to preserve inter-individual variability of interest.

Amyloid-PET images were processed using standardized ADNI pipelines. Standardized uptake value ratios (SUVRs) were computed by normalizing tracer uptake to the whole cerebellum as a reference region. Regional SUVR values were extracted using the Desikan–Killiany (DK) atlas ^37^ for cortical regions and the aseg atlas ^38^ for subcortical regions and were subsequently used for downstream statistical analyses. Detailed preprocessing procedures for PET imaging are provided in the ADNI_UCBerkeley_AmyloidPET_Methods_v3_2025-06-30.pdf document, available through the ADNI database.

### 4.3 Normative modeling of regional gray matter atrophy

A normative modeling approach was applied to estimate individualized deviations in regional GMV (Figure 1A). Specifically, regional GMV was first modelled in the healthy controls (CN Aβ− group) using the general linear model, with age, sex, handedness, education and total intracranial volume as covariates. The model was used to estimate expected GMV for each region based on an individual’s demographic characteristics. For patients in the Aβ+ CN-NC, Aβ+ CN-C, Aβ+ MCI-NC, Aβ+ MCI-C, and Aβ+ AD, individual covariate values were entered into the corresponding regression models to obtain predicted regional GMV. W-scores were then computed for each region as the difference between the predicted and observed GMV, divided by the standard error of the regression model estimated in the CN Aβ− group ^39^. Higher W-scores indicate greater deviation from the normative reference, corresponding to more pronounced gray matter atrophy. Positive and negative W-scores indicate deviations above and below the normative ranges, respectively. Given that the present study specifically focused on gray matter atrophy, only positive W-scores were included for subsequent analyses. This W-score framework enables the construction of individualized maps of gray matter atrophy while accounting for demographic and anatomical covariates that influence GMV ^39–41^.

### 4.4 Multiscale connectome-based node-neighbor relationships

Multiscale cortical connectomes (Figure 1B), including structural connectivity (SC), gene co-expression (GC), neurotransmitter receptor similarity (RS), laminar similarity (LS), metabolic connectivity (MC), electrophysiological connectivity (EC), temporal similarity (TS) and functional connectivity (FC), were obtained from a previously published study ^17^. Detailed construction procedures are described in Supplementary Methods 1. Edge weights in the SC matrix were rescaled to the [0, 1] interval, while all other connectomes were normalized using the Fisher’s r-to-z transform and restricted to positive values. The rescaled structural connectivity matrix was used as the weighted structural connectivity (wSC), whereas a binarized version, in which edges with nonzero weights were set to 1 and all other entries to 0, was used as the binary structural connectivity (bSC). To assess the influence of FC sparsification, we retained the strongest 90%, 80%, 70%, 60%, 50%, 40%, 30%, 20%, 10%, and 5% of connections in the FC matrix, yielding networks denoted FC90 to FC5, respectively.

Neighbors of a given region were defined as regions connected to it by a nonzero edge weight (Figure 1B). For each cortical region *i*, the collective atrophy of its neighborhood (*p_j_*) was computed as the weighted average W-score of all connected neighboring regions ^42–44^:

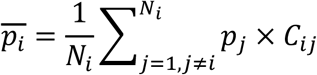

where *N*_i_ denotes the number of connected neighbors of region *i* (i.e., node degree), *p_j_* represents the W-score value observed in the *j*-th neighboring region, and *C_ij_* is the connectivity strength between regions *i* and *j*. Under this formulation, neighbors with stronger connections contributed more strongly to the estimation of neighborhood-level atrophy associated with region *i*, which meant stronger connections contribute more to neighborhood atrophy. Spearman’s correlation was used to assess the association between node-level deviation prevalence values and their corresponding mean neighbor prevalence values across cortical regions.

### 4.5 Null models

For each group, regional W-scores were averaged across participants to derive a mean atrophy map, from which group-level node–neighbor relationships were calculated. Statistical significance was evaluated using three null models: Null-spin, Null-smash, and Null-rewired (Figure 1B). Null-spin and Null-smash tested whether the observed relationships exceeded those expected from surrogate W-score maps with approximately preserved spatial autocorrelation. The two models differ in the way in which they account for the spatial structure present in the data. First, Null-spin used a spin-test ^45^ implemented in neuromaps ^46^ to rotate the cortical map 5,000 times to generate surrogate regional maps. The rotation was applied to one hemisphere and then mirrored for the other hemisphere. Second, Null-smash used a parametric model to capture the spatial structure. Specifically, it relies on spatial variogram modeling to generate 5,000 random spatial maps with a similar spatial autocorrelation to the observed atrophy map, as implemented in the toolbox BrainSMASH ^47^. Third, Null-rewired randomized each connectome using netneurotools ^48^, preserving its degree sequence and overall edge-weight distribution and approximately preserving its edge-length distribution and weight–length relationship ^49^. Rewiring used 10 distance bins to generate 5000 rewired networks. Across all three null models, the surrogate values were used for inference on the observed performance metrics, with p-values quantified as the fraction of null values exceeding the observed value. FDR correction was applied separately for each null model across all groups and all tested architectures.

### 4.6 Subject-level comparisons of node–neighbor relationships across connectomes

Differences in node–neighbor relationships across connectomes were examined by comparing each connectome-specific measure to that derived from the Euclidean distance network. Analyses were conducted separately within each group. Subject-level node–neighbor relationships were Fisher’s z-transformed. For each connectome, a paired-samples t-test was performed to estimate whether node–neighbor relationships were significantly higher than those based on Euclidean distance, with FDR correction applied to account for multiple comparisons across connectomes.

### 4.7 Longitudinal changes in node–neighbor relationship

Longitudinal changes in node–neighbor relationships were examined separately for each clinical group using linear mixed-effects models. Fisher r-to-z transform node–neighbor relationships were modelled as a function of time since baseline, connectome type and their interaction, with baseline age, sex, education years and handedness included as covariates. Participant-specific random intercepts and slopes for time accounted for differences in baseline values and rates of change. The time × connectome interaction was used to assess whether longitudinal changes differed among connectomes.

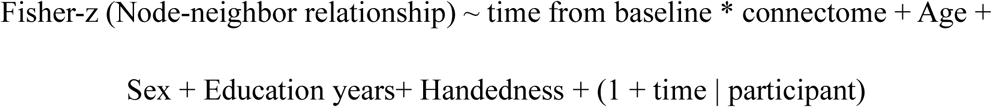

### 4.8 Network diffusion modeling

Network diffusion modeling (NDM)^7,43^ was independently initialized using each of the 400 cortical regions as a seed (Figure 1C). At each initialization, the seed region was assigned an initial pathology value of 1, with all other regions set to 0. The diffusion process was then simulated over model time steps ranging from t = 0 to 50, using a fixed diffusion constant (a = 1). Formally, the NDM describes the evolution of pathology over time as:

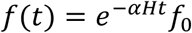

where f(t) denotes the regional pathology distribution at diffusion time t, *H* is the Laplacian of the weighted connectivity matrix, and *f₀* represents the initial seeding configuration.

For each participant and connectome, each brain region was used in turn as a seed in the NDM. At each simulated time point, we calculated the Pearson correlation between the simulated atrophy pattern and the participant’s observed baseline pattern, retaining the maximum correlation (*r*_max)_ for each seed. Seed-specific *r*_max v_alues were then compared with null distributions generated *r*_max u_sing the Null-spin and Null-smash models. A seed region was defined as a candidate epicenter if its *r*_max s_ignificantly greater than that of a spatially constrained null model (i.e., *P*_Null-spin_ < 0.05 or *P*_Null-smash_ < 0.05).

Follow-up visits were defined as assessments conducted at 6, 12, 24 or 36 months after baseline, allowing a ±2-month window around each time point. At each follow-up visit, NDM patterns simulated from the candidate epicenters were compared with the observed GMV alteration pattern. Longitudinal model prediction performance was defined as the highest Pearson correlation across candidate epicenters and simulated time points. Prediction performance was compared between connectomes using paired *t*-tests on data from participants assessed at the same follow-up time point. Because the number of candidate epicenters differed across connectomes, and a maximum taken over more candidates is inflated relative to one taken over fewer, we recorded the number of candidate epicenters per participant and connectome and repeated the comparisons using a fixed number of top ranked epicenters for every connectome.

### 4.9 Validation analysis

To assess the reproducibility of the findings, we repeated the analyses in the independent OASIS-3 cohort. T1-weighted MRI scans were processed to obtain GMV for the same 400 cortical regions used in the discovery cohort, and regional GMV measures were harmonized across imaging sites using ComBat. Individual regional atrophy W-scores were estimated using Aβ− CN participants in OASIS-3 as the normative reference. Node–neighbor relationships were calculated for each connectome and evaluated against Null-spin, Null-smash and Null-rewired distributions. At the participant level, connectome-specific relationships were compared with the Euclidean-distance benchmark using paired tests and were also compared across clinical groups.

## Data Availability

Data used in this study are available from the Alzheimer’s Disease Neuroimaging Initiative (ADNI; https://adni.loni.usc.edu) and Open Access Series of Imaging Studies (OASIS-3; https://www.oasis-brains.org) repositories. The group-level multiscale connectomes are publicly available from Hansen et al. ^17^.

## Code Availability

Cortical maps were visualized using the ggseg package ^50^. Analysis code supporting the findings of this study is available at https://github.com/kele-gao/AD-multiscale-connectome-atrophy.

## Acknowledgements

We would like to thank the anonymous reviewers for their constructive comments, that improved the quality of this work substantially.

## Author contributions

K.G. contributed conceptualization, methodology, software, visualization, writing—original draft, and writing—review & editing. Y.S. contributed software. J.B. contributed software. D.Y. contributed writing—review & editing. B.B. B. contributed writing—review & editing and supervision. P.W. contributed writing—review & editing and supervision.

