## Supplementary file for "Multiscale connectomes reveal stage-dependent organization of cortical atrophy in Alzheimer’s disease"

### Supplementary Methods

Structural connectivity (SC) represented anatomical coupling derived from diffusion-weighted imaging (DWI) data acquired from 326 unrelated participants (age range: 22–35 years; 145 males) in the Human Connectome Project (HCP; S900 release) [1]. We examined both weighted (wSC) and binary (bSC) representations of SC. The bSC matrix encoded the presence or absence of structural connections as 1 or 0, respectively. Functional connectivity (FC) was computed between blood-oxygen-level-dependent (BOLD) time series derived from resting-state functional magnetic resonance imaging (fMRI) data acquired from the same 326 unrelated participants (age range: 22–35 years; 145 males) in the HCP (S900 release) [1]. Temporal similarity (TS) quantified higher-order dynamic similarity between cortical regions by correlating feature representations of BOLD time series, capturing temporal dependencies beyond conventional Pearson correlation used in FC. TS was derived from the same resting-state fMRI data used to construct FC. Gene co-expression (GC) quantified transcriptional similarity between cortical regions based on expression profiles of more than 8,000 genes from the Allen Human Brain Atlas (AHBA) [2], derived from six postmortem brains (one female; age range: 24.0–57.0 years; mean ± SD: 42.5 ± 13.38 years). Neurotransmitter receptor similarity (RS) captured inter-regional similarity in protein density across 18 neurotransmitter receptors and transporters spanning nine neurotransmitter systems: dopamine (D1, D2, DAT), norepinephrine (NET), serotonin (5-HT1A, 5-HT1B, 5-HT2, 5-HT4, 5-HT6, 5-HTT), acetylcholine (α4β2, M1, VAChT), glutamate (mGluR5), γ-aminobutyric acid (GABAA), histamine (H3), cannabinoid (CB1), and opioid (MOR). PET tracer images for 18 neurotransmitter receptors and transporters were obtained from a previous study[3] and neuromaps [4]. Laminar similarity (LS) was derived from correlations of cell-staining intensity profiles across cortical layers using the BigBrain data [5], obtained from the postmortem brain of a male (age > 60). Metabolic connectivity (MC) was defined as the correlation of dynamic $^{18}$F-fluorodeoxyglucose positron emission tomography (FDG-PET) time series obtained from 26 healthy participants (77% female; age range: 18–23 years) [6], reflecting inter-regional similarity in glucose uptake. Electrophysiological connectivity (EC) was derived from resting-state magnetoencephalography (MEG) data acquired from 33 unrelated healthy young adults (age range: 22–35 years) in the HCP (S900 release) [1] and defined as the first principal component of connectivity patterns across six canonical frequency bands. To further examine the influence of spatial proximity, the reciprocal of the Euclidean distance between regional centroids was calculated to represent the physical closeness of regions. Detailed construction procedures for each connectome have been described previously [7]. Additionally, all connectomes were constructed at the regional level using the Schaefer atlas with 400 cortical regions [8].

**Supplementary Figures**

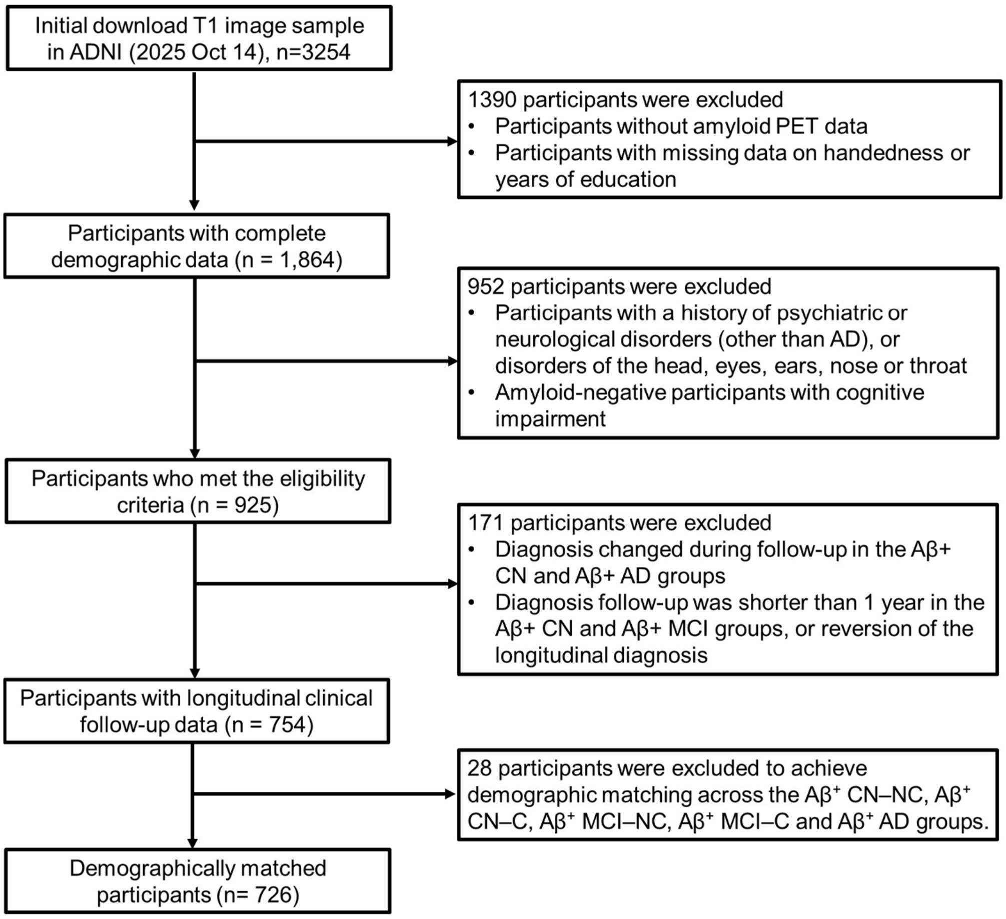


**Supplementary Figure 1. Participants selection flowchart for the discovery cohort.** ADNI: Alzheimer's Disease Neuroimaging Initiative; Aβ+ CN-NC: Aβ-positive cognitively normal non-converters; Aβ+ CN-C: Aβ-positive cognitively normal converters; Aβ+ MCI-NC: Aβ-positive mild cognitive impairment non-converters; Aβ+ MCI-C: Aβ-positive mild cognitive impairment converters; Aβ+ AD: Aβ-positive dementia

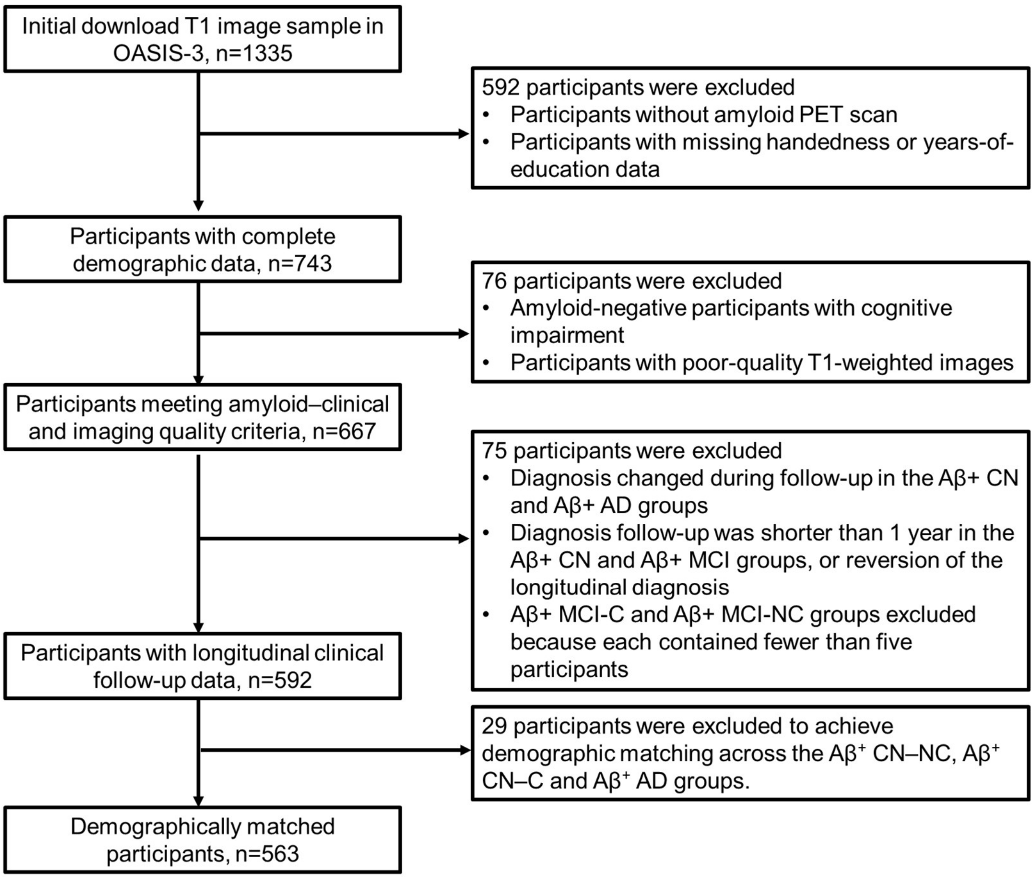


**Supplementary Figure 2. Participants selection flowchart for the validation cohort.** OASIS-3: Open Access Series of Imaging Studies; Aβ+ CN-NC: Aβ-positive cognitively normal non-converters; Aβ+ CN-C: Aβ-positive cognitively normal converters; Aβ+ MCI-NC: Aβ-positive mild cognitive impairment non-converters; Aβ+ MCI-C: Aβ-positive mild cognitive impairment converters; Aβ+ AD: Aβ-positive dementia

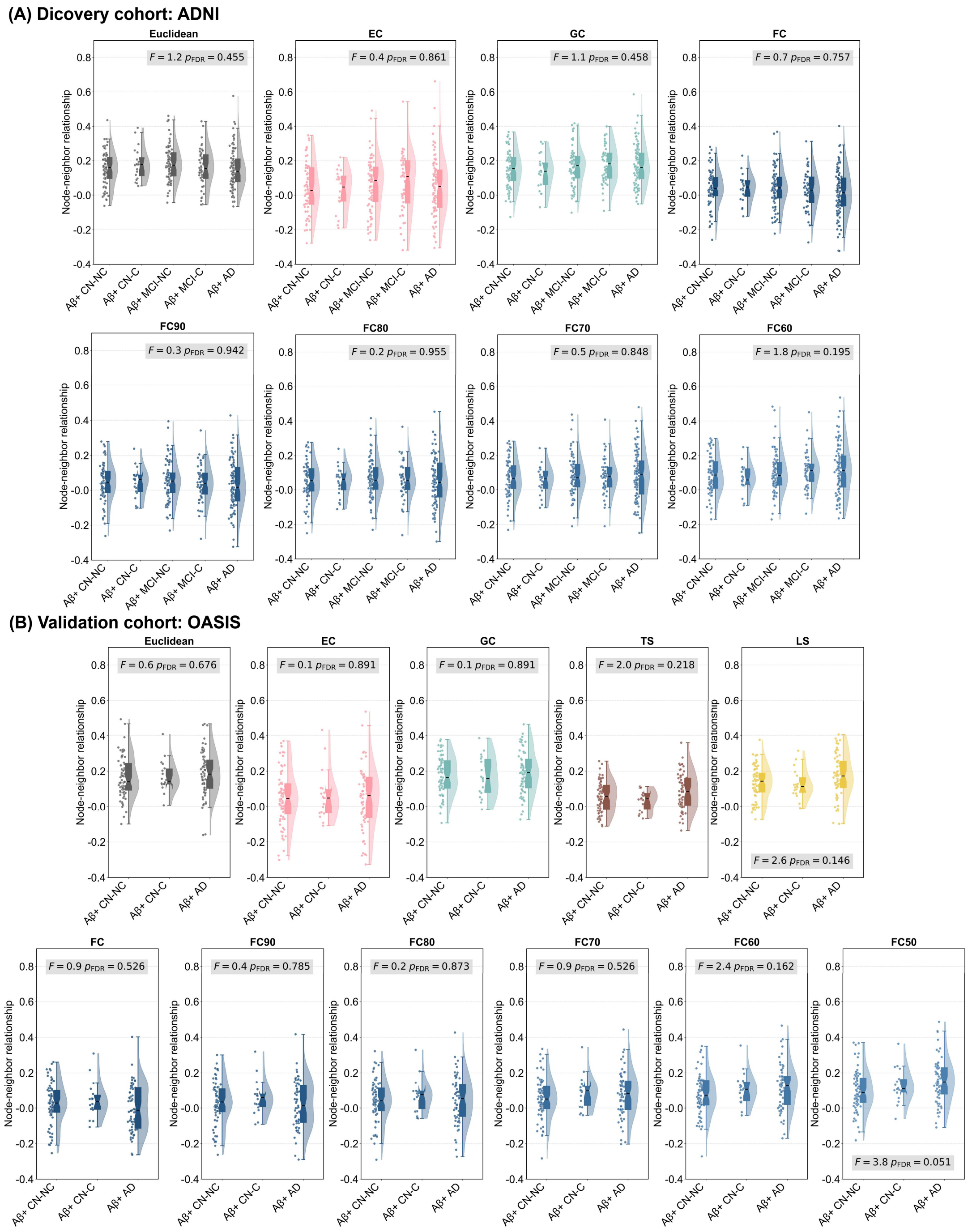


**Supplementary Figure 3. Comparisons of node–neighbor relationships across groups.** Statistical results from analysis of covariance (ANCOVA) are displayed within each panel. ADNI: Alzheimer's Disease Neuroimaging Initiative; OASIS-3: Open Access Series of Imaging Studies; Aβ+ CN-NC: Aβ-positive cognitively normal non-converters; Aβ+ CN-C: Aβ-positive cognitively normal converters; Aβ+ MCI-NC: Aβ-positive mild cognitive impairment non-converters; Aβ+ MCI-C: Aβ-positive mild cognitive impairment converters; Aβ+ AD: Aβ-positive dementia; EC: electrophysiological connectivity; GC: gene co-expression; FC: functional connectivity. FC90–FC60: retaining the strongest 90%–60% of FC connections.

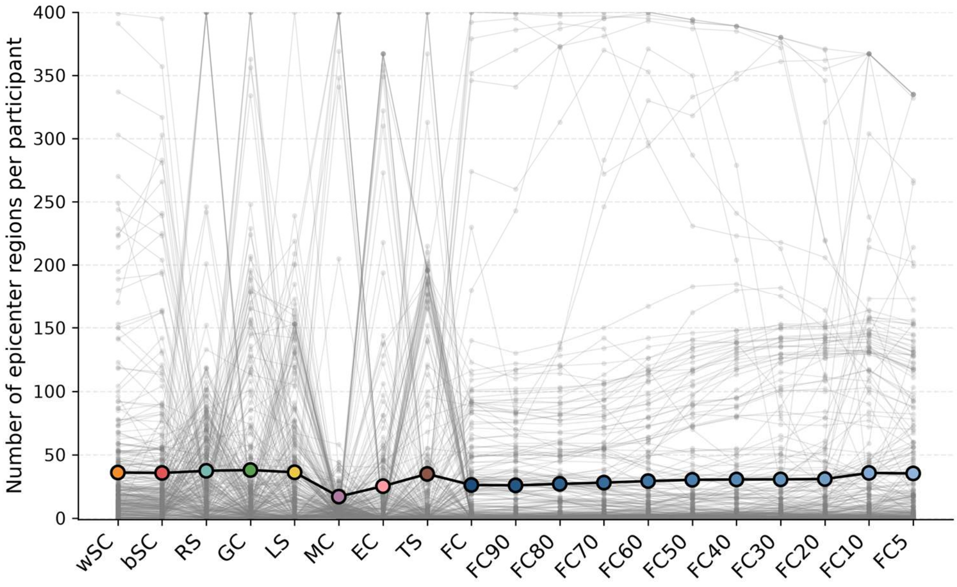


**Supplementary Figure 4. Number of candidate epicenter regions identified for each connectome.** Each grey line connects the numbers of candidate epicenter regions identified for an individual participant across connectomes; grey points represent individual values. Colored points show the mean number of candidate epicenters for each connectome. bSC: binary structural connectivity; wSC: weighted structural connectivity; GC: gene co-expression; RS: neurotransmitter receptor similarity; LS: laminar similarity; MC: metabolic connectivity; EC: electrophysiological connectivity; TS: temporal similarity; FC: functional connectivity. FC90–FC5: retaining the strongest 90%–5% of FC connections.

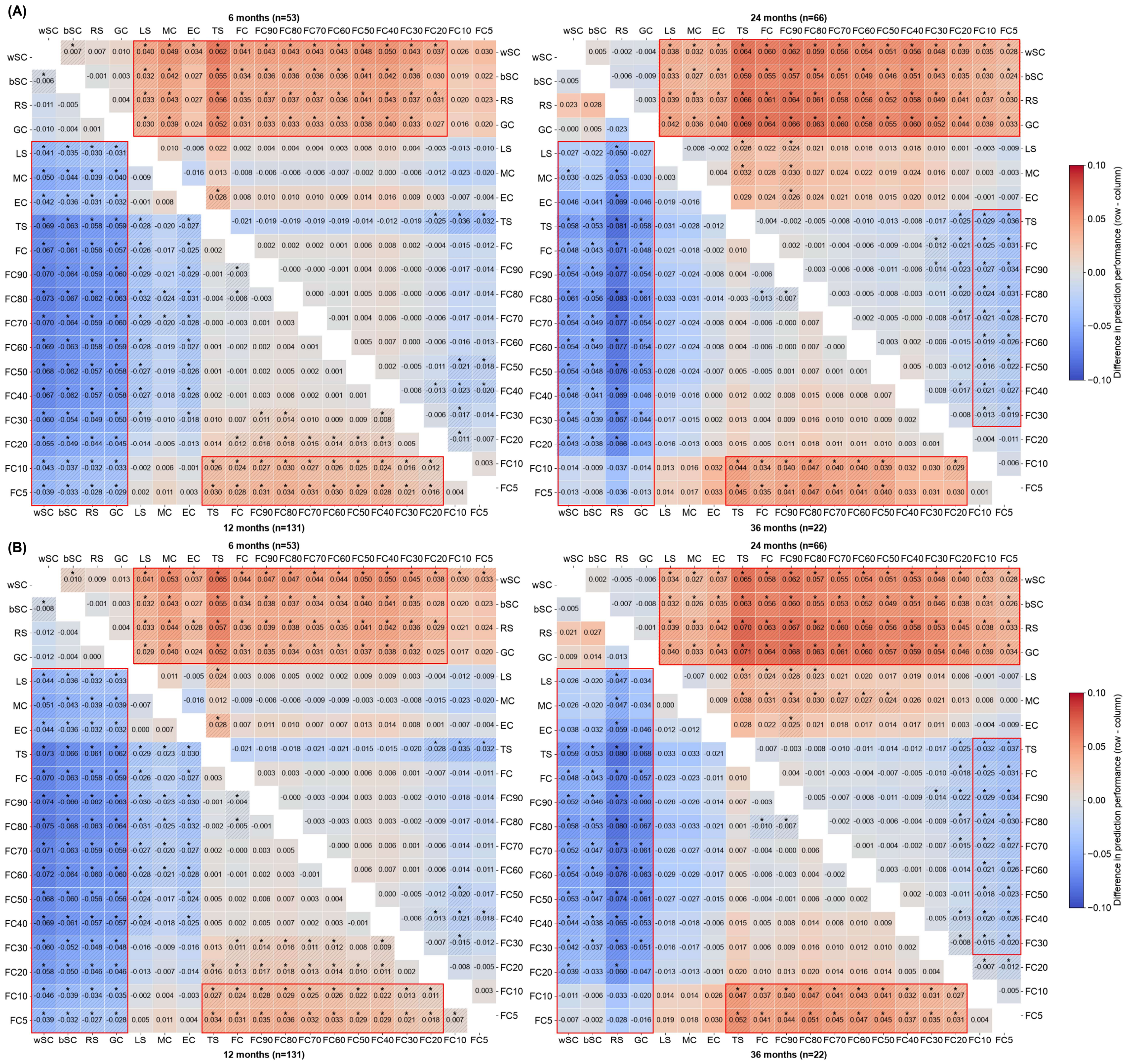


**Supplementary Figure 5. Pairwise comparisons of network diffusion model (NDM) prediction performance at 6, 12, 24 and 36 months.** (A) and (B) show results obtained by retaining the five and ten highest-ranked seed regions, respectively, as candidate epicenters. Cell values represent the mean difference in NDM prediction performance between the connectome indicated by the row and column labels. Asterisks and diagonal hatching mark significant differences ($P_{FDR} < 0.05$). The n range shown for each follow-up interval indicates the minimum and maximum numbers of participants with prediction estimates for both connectomes in a pairwise comparison.

Each participant contributed at most one visit within the ±2-month window around the target month; comparisons with fewer than 10 paired participants are shown as N.A. bSC: binary structural connectivity; wSC: weighted structural connectivity; GC: gene co-expression; RS: neurotransmitter receptor similarity; LS: laminar similarity; MC: metabolic connectivity; EC: electrophysiological connectivity; TS: temporal similarity; FC: functional connectivity. FC90–FC5: retaining the strongest 90%–5% of FC connections.

### Supplementary Tables

**Supplementary Table 1. Demographic and clinical characteristics of the Aβ− CN group.**

| | | Discovery cohort: ADNI Aβ- CN (n=418) | Validation cohort: OASIS-3 Aβ- CN (n=411) |
|---|---|---|---|
| Age (mean years ± SD) | | 67.9±6.5 | 65.2±8.6 |
| Women n(%) | | 270(64.6%) | 266(64.7%) |
| Education years (mean years ± SD) | | 16.6±2.3 | 15.9±2.4 |
| Handedness, n(%) | right | 371(88.8%) | 373(90.8%) |
| | left | 47(11.2%) | 38(9.2%) |
| CDR n(%) | 0 | 416(99.5%) | 411(100%) |
| | 0.5 | 2(0.5%) | 0(0%) |
| | 1 | 0(0%) | 0(0%) |
| MMSE scores (mean ± SD) | | 29.1±1.2 | 29.2±1.1 |

ADNI: Alzheimer's Disease Neuroimaging Initiative; OASIS-3: Open Access Series of Imaging Studies;

**Supplementary Table 2. Diagnostic follow-up and time to clinical progression across the study groups in Discovery cohort**

| | Aβ- CN | Aβ+ CN-NC | Aβ+ CN-C | Aβ+ MCI-NC | Aβ+ MCI-C | Aβ+ AD |
|---|---|---|---|---|---|---|
| Number of diagnostic assessments, median (IQR) | 3 (2–4) | 4 (3–6) | 5 (4–6.2) | 4 (3–6) | 5 (4–6) | 3 (2–4) |
| Diagnostic follow-up duration, years, median (IQR) | 2.1 (0.2–6.7) | 6.3 (3.1–7.1) | 5.3 (3.4–7.3) | 2.6 (1.2–4.7) | 3.4 (2.4–5) | 1.1 (0.6–2.1) |
| Diagnostic follow-up duration, years, minimum–maximum | 0–14 | 1–11.7 | 1.2–13.2 | 1–14 | 1–10.6 | 0–7 |
| Diagnostic follow-up duration, years, mean±SD | 3.3±3.4 | 5.2±2.7 | 5.7±3.3 | 3.2±2.3 | 4±2.2 | 1.5±1.5 |
| Time to first qualifying progression, years, median (IQR) | - | - | 4.1 (1.4–6.2) | - | 2.3 (1.4–3.4) | - |

Aβ+ CN-C: Aβ-positive cognitively normal converters; Aβ+ MCI-NC: Aβ-positive mild cognitive impairment non-converters; Aβ+ MCI-C: Aβ-positive mild cognitive impairment converters; Aβ+ AD: Aβ-positive dementia
